# Domains of Frailty as Early Risk Factors for Alzheimer’s Disease – Genetic and Causal Evidence

**DOI:** 10.1101/2025.09.17.25336027

**Authors:** Jonny P Flint, Isabelle F Foote, David D Ward, Tom C Russ, Alan Marshall, Md Rezanur Rahman, Simon R Cox, Michelle Luciano, Michelle K Lupton

## Abstract

**Background:** Alzheimer’s disease (AD) is a leading cause of disability and dependency in older adults. Frailty is a phenotypic risk factor for AD, but its causal role remains unclear – partly due to reliance on composite scores that may obscure distinct mechanisms.

**Methods:** We applied a Mendelian randomisation (MR) framework using two large GWAS of clinically diagnosed AD. Frailty exposures were derived from a multivariate Genomic SEM model comprising one general factor and six domain-specific domains. Bidirectional univariable MR (UVMR) estimated total effects on AD, while multivariable MR (MVMR) adjusted for educational attainment, household income, and longevity.

**Results:** The general frailty factor showed no evidence of a causal effect on AD. The unhealthy lifestyle frailty domain increased AD risk in UVMR (OR = 6.60, 95% CI: 2.36-18.50, q = 0.021 and OR = 3.12, 95% CI: 1.57-6.19, q = 0.001), but these effects attenuated substantially in MVMR, with betas reduced by ∼66-94% and ORs falling to ∼1.1-1.5 (nonsignificant), consistent with overlap with socioeconomic pathways. The disability frailty domain was nominally protective in UVMR, but positively associated with AD in MVMR (OR = 1.69, 95% CI: 1.21-2.36, q = 0.007). The poorer cognition frailty domain increased AD risk in MVMR when adjusted for education and income (OR = 1.55, 95% CI: 1.12-2.15, q = 0.025), though this effect attenuated by ∼37% with further adjustment for longevity. Sensitivity analyses indicated that this cognition-AD signal was partly driven by SNPs in SPI1, a well-established AD risk locus, consistent with pleiotropic overlap rather than an independent domain. No reverse effects were observed.

**Conclusions:** Frailty comprises genetically distinct domains with differential causal relevance for AD. These findings highlight disability-related frailty as a potential causal contributor to AD, whereas cognitive-related frailty is likely predominantly driven by shared aetiology, underscoring the need to move beyond composite frailty indices when evaluating dementia risk.

## Introduction

Alzheimer’s disease (AD) is a heterogeneous neurodegenerative disorder and a leading cause of disability in older adults ^[1]^. In 2020, an estimated 60 million people worldwide were living with AD – a figure projected to rise to 153 million by 2050 ^[2]^. Identifying modifiable risk factors is a key public health priority. The decrease in age-specific incidence of dementia in high-income countries has led to an increased focus on dementia prevention, with the most recent Lancet Commission report on dementia prevention, intervention and care estimating that up to 45% of all-cause dementia might be prevented through reducing modifiable risk factors ^[3]^. However, much of this evidence derives from observational studies, which cannot reliably distinguish causal risk factors from those associated through reverse causation or confounding. Modifying risk factors will only reduce AD prevalence if those factors are truly on the causal pathway.

Mendelian Randomisation (MR) is a method that leverages genetic variation to infer causal relationships between exposures and outcomes ^[4]^. It is based on the principle that genetic variants are randomly assorted at conception, mirroring the randomisation process in a controlled trial. By using genetic variants robustly associated with an exposure as instrumental variables, MR can help disentangle whether an observed association with an outcome reflects a true causal effect rather than confounding or reverse causation. In contrast to traditional observational studies, which are vulnerable to biases arising from lifestyle, socioeconomic, or environmental differences, MR offers a quasi-experimental framework that is less susceptible to such confounding. This has spurred growing interest in identifying risk factors that are not only causally linked to AD but may also be more readily modifiable across the life course ^[5]^.

Frailty represents one such candidate – conceptualised as a multidimensional syndrome reflecting reduced physiological reserve and increased vulnerability to stressors ^[6]^. Frailty has consistently been associated with increased risk of cognitive decline and AD ^[6,7]^. In a recent phenotypic analysis of ∼30,000 participants across four UK and US cohorts, it was found that frailty levels were elevated 8 to 20 years prior to AD onset, with the rate of decline in health and function accelerated markedly in the 4 to 9 years preceding diagnosis ^[8]^. Importantly, frailty predicted incident AD even when measured well before the prodromal phase, positioning it as a potential early risk signal of neurodegenerative vulnerability. Frailty may actively contribute to neurodegeneration by accelerating biological ageing, increasing vulnerability to comorbidities, and reducing systemic resilience ^[9,10]^.

Despite strong phenotypic evidence, the biological mechanisms linking frailty to AD remain poorly understood. Most genetic studies to date have relied on composite frailty scores – including the Frailty Index ^[11]^, Frailty Phenotype ^[12]^ and the Hospital Frailty Risk Score ^[13]^ – which aggregate heterogeneous health indicators into a single summary measure. While clinically useful, such aggregated scores may obscure distinct biological pathways and dilute genetic signal ^[14,15]^. We recently published the first multivariate GWAS of frailty, which applied Genomic Structural Equation Modelling (Genomic SEM) to model the latent pattern of shared genetics between 30 frailty-related traits ^[15]^. This modelling approach identified a complex genetic architecture comprising a general frailty factor and six additional factors capturing distinct dimensions of frailty that were uncorrelated with the general factor, suggesting separable biological pathways. Importantly, these domains displayed different patterns of genetic correlation with age-related health outcomes. For example, whilst the general frailty domain was not genetically correlated to AD, the frailty domain poorer cognition was significantly associated with AD, indicating that there may be specific genetically-informed sub-domains of frailty that could be the main drivers of the observational association that is seen between frailty and AD. A previous MR study using composite frailty indices also reported null associations with AD ^[16]^, but causal relationships between the more nuanced sub-domains of frailty have not previously been explored.

Therefore, we used a bidirectional two-stage MR approach to investigate whether genetically derived frailty sub-domains have a causal impact on AD. First, we performed univariable MR (UVMR) to estimate the independent effects of general frailty and each of the six domain-specific factors on AD risk, using summary statistics from two large GWAS of clinically diagnosed AD. We focused on the studies by Kunkle et al. (2019) and Wightman et al. (2021), which together provide complementary strengths: rigorous diagnostic ascertainment with close age-matching (Kunkle et al. 2019) and the largest case-control sample to date through integration of biobank and clinical cohorts (Wightman et al. 2021, excluding proxy cases) ^[17,18]^. We then used multivariable MR (MVMR) to assess whether frailty exerts a direct causal effect on AD, or whether its influence is mediated or confounded by socioeconomic factors, lifespan-related selection, or other indirect pathways ^[19]^. Our study provides the first comprehensive MR investigation of frailty and AD by leveraging a multivariate genetic framework to offer a more nuanced understanding of how specific aspects of frailty may contribute to AD risk, whilst adhering to the STROBE-MR reporting guidelines to ensure methodological transparency and reproducibility.

## Methods

### GWAS summary statistics

#### Frailty exposures

Frailty exposures were defined using summary statistics from our previously published multivariate GWAS analysis of 30 frailty-related traits using Genomic SEM ^[15]^. This model identified seven latent frailty constructs based on a broad set of clinical, behavioural, and psychosocial indicators (see the original paper for detailed descriptions of the individual indicators ^[15]^). These included a general frailty factor (GF) of shared genetics between all frailty deficits (which is most comparable to prior aggregate scores) and six residual factors (F1-F6) of shared genetics between different subsets of deficits that were uncorrelated with the GF. In brief, these residual factors can be viewed as frailty genetic domains linked to limited social support (F1), unhealthy lifestyle (F2), multimorbidity (F3), metabolic problems (F4), poorer cognition (F5) and disability (F6) (summarised in Table 1 and Figure 1).

**Table 1.**
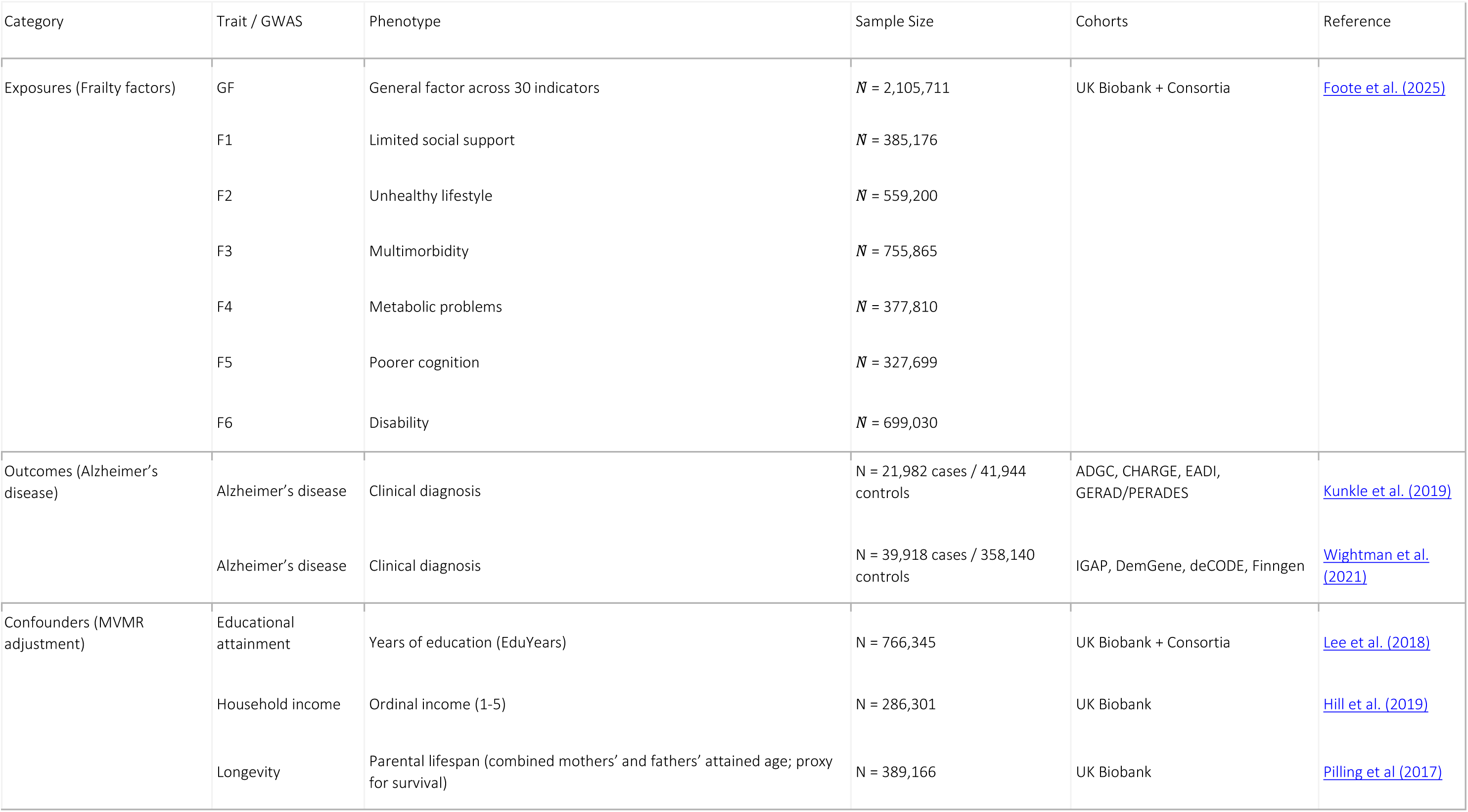
All genome-wide association studies (GWAS) used as exposures in downstream analyses, including Alzheimer’s disease and frailty-related traits. For frailty, we include the general latent factor and six orthogonal residual factors (F1-F6), each representing distinct domains – sample sizes reflect effective N (*N̂*) as estimated by Genomic SEM. UK Biobank was the primary source of data for most frailty indicators, but several traits included data from large consortia. Further details on phenotypes, measurement, ancestry, and cohorts can be found in Supplementary Table 1.

**Figure 1.**
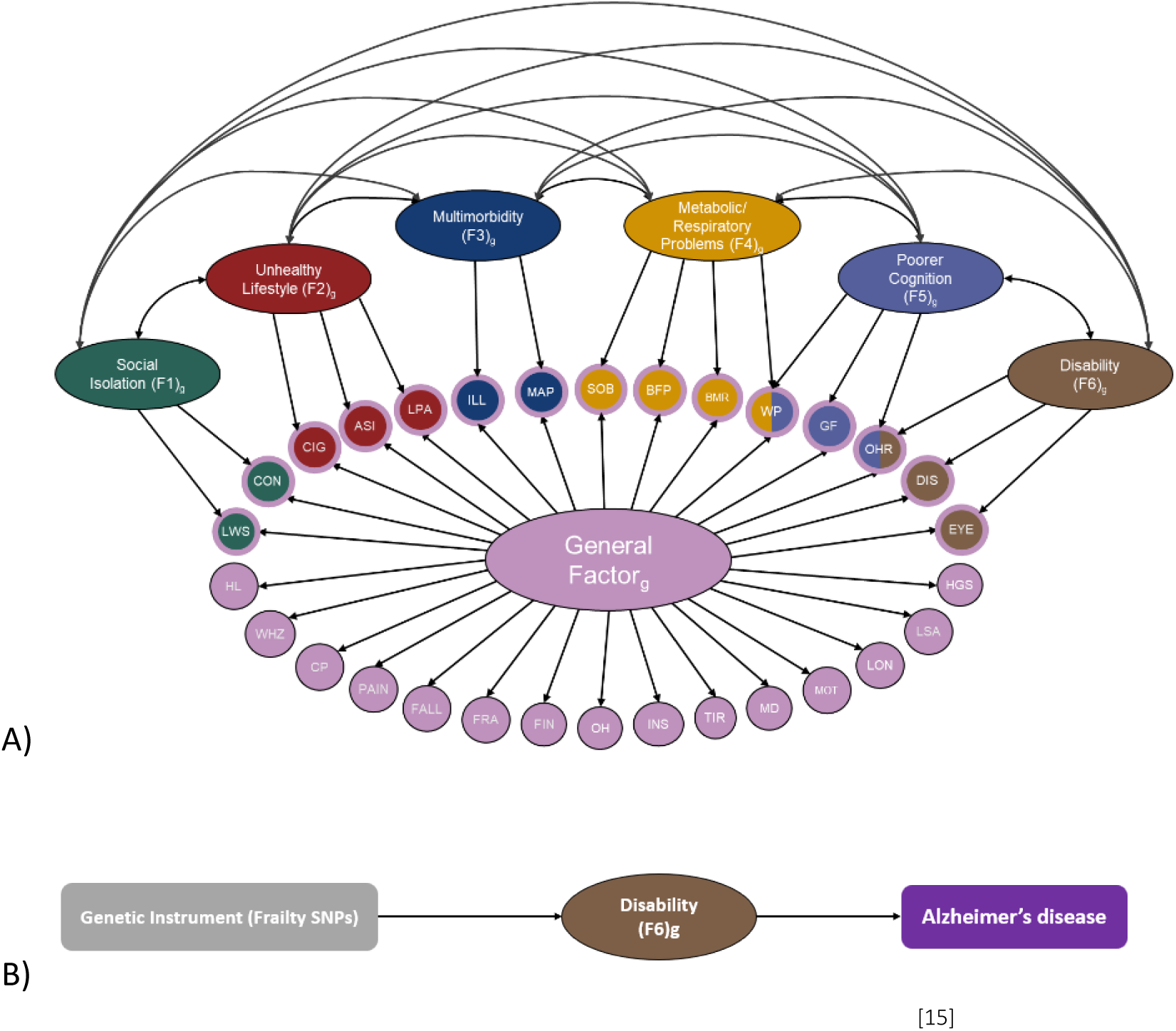
UVMR schematics. Panel A: Path diagram of the multivariate frailty model (adapted from Foote et al, 2025), shown without factor loadings to highlight the overall structure (please refer to the original paper for the full statistical properties of the model). The large oval represents the general factor of frailty (GF), which is orthogonal to the six domain-specific domains (F1-F6; medium-size ovals). The small circles represent the 30 measured frailty indicators. Acronyms for indicators are defined as follows: ASI, pulse wave arterial stiffness index; BFP, body fat percentage; BMR, basal metabolic rate; CIG, number of cigarettes smoked per day; CON, unable to confide; CP, chest pain; DIS, long-standing illness, disability or infirmity; EYE, eye disorder or problem; FALL, number of falls in past year; FIN, financial difficulties; FRA, fracture in last 5 years; GF, low fluid intelligence score; HGS, low hand grip strength; HL, age related hearing loss; ILL, number of non-cancer illnesses; INS, insomnia; LON, loneliness or isolation; LPA, physical inactivity; LSA, low social or leisure activity; LWS, not living with spouse or partner; MAP, mean arterial pressure; MDD, major depressive disorder; MOT, feelings of unenthusiasm or disinterest; OH, poor oral health; OHR, poorer overall health rating; PAIN, pain experienced in the past month; SOB, shortness of breath when walking on flat ground; TIR, tiredness or lethargy; WHZ, wheezing or whistling in chest in the past year; WP, slow walking pace. Panel B: Simplified path diagram illustrating the UVMR framework, shown with a single frailty domain (‘Disability’ as an example exposure) and Alzheimer’s disease (AD) as the outcome for clarity. This analysis was performed separately for the GF and each of the six domain-specific frailty domains (F1-F6). The directly measured phenotypes (that is the SNP effects and the univariate AD outcome) are depicted as rectangles and the latent frailty measures are depicted as an oval.

#### Alzheimer’s disease outcomes

We analysed two large-scale GWAS of clinically diagnosed Alzheimer’s disease: Kunkle et al. 2019 (21,982 cases, 41,944 controls) and Wightman et al. 2021 (39,918 cases, 358,140 controls after excluding proxy-based definitions). AD Kunkle draws on 46 datasets with clinically or autopsy-confirmed diagnoses and closely age-matched cases and controls (mean difference 0.4 years), minimising age-related confounding. AD Wightman, in which we excluded proxy cases, is the largest and most recent clinically defined case control GWAS, it integrates biobank and clinical cohorts with ICD-based ascertainment, providing substantial power, but with limited reporting on age-matching and heavy reliance on population-based cohorts such as deCODE and UK Biobank. This focus on the two largest and most recent non-proxy, clinically defined GWAS prioritised phenotypic specificity while still providing complementary strengths: diagnostic precision in Kunkle and statistical power in Wightman. As highlighted in AD genetic research, larger GWAS that include proxy-based or less strictly defined phenotypes may introduce noise; by focusing on clinically diagnosed cases, we prioritised specificity over size to ensure reliable causal inference ^[20,21]^.

#### Confounders: Education attainment, Household Income and Longevity

To assess whether associations between frailty and AD are mediated or confounded by socioeconomic factors and lifespan-related selection, we included educational attainment (EA), household income, and genetic liability to longevity as additional exposures in multivariable Mendelian randomisation (MVMR) analyses ^[20–22]^; details for each confounder are listed in Table 1. EA summary statistics were based on a GWAS of 726,808 individuals of European ancestry ^[22]^, and GWAS data for household income were derived from 286,301 UK Biobank participants ^[23]^. Longevity summary statistics were obtained from a large GWAS of parental lifespan in 389,166 UK Biobank participants ^[24]^. We selected this study (Pilling et al. 2017) due to its large sample size, which provides strong instruments for survival-related genetic liability. All three traits are established correlates of frailty and cognitive function; EA and income have been shown in previous MR studies to causally influence AD risk ^[4,5]^, while longevity captures genetic influences on survival that can contribute to survival bias and right censoring in dementia research ^[25]^. Including EA, income, and longevity in MVMR allows us to determine whether frailty domains exert direct effects on AD, or whether their effects are better explained by socioeconomic pathways or selective survival processes.

### Mendelian Randomisation Analyses

Mendelian randomisation (MR) is a statistical method that uses genetic variants as instrumental variables (IVs) to test for causal relationships between an exposure and an outcome ^[26,27]^. For a genetic variant to be considered a valid instrument, three core assumptions must be satisfied: (1) the variant is robustly associated with the exposure (relevance), (2) it affects the outcome only via the exposure (exclusion restriction), and (3) it is not associated with confounders of the exposure-outcome relationship (independence). This paper followed STROBE-MR guidelines ^[28]^, as documented in supplementary materials. All MR analyses were conducted in R (version 4.3.1) ^[29]^ using the most recent versions of the TwoSampleMR ^[30]^, MVMR ^[31]^, and MRPRESSO packages ^[32]^. Genome-wide significant SNPs (p < 5 × 10⁻⁸) were selected as instruments and clumped for linkage disequilibrium (LD) using the clump_data() function in TwoSampleMR (r² < 0.001, window = 10,000 kb), based on the 1000 Genomes European reference panel. Variants within the APOE locus (chr19:44, 909, 03945, 912, 650; hg19), including a 500 kb flanking region, were excluded from all analyses to avoid bias due to the large effect size and extended linkage disequilibrium of this region. For the multivariable models, we combined all genome-wide significant SNPs identified for any of the included exposures into a single instrument set (that is the union of exposure-specific instruments, rather than restricting to overlapping SNPs).

#### Univariable Mendelian Randomisation (UVMR)

We conducted univariable Mendelian randomisation (UVMR) analyses using the inverse variance weighted (IVW) method as the primary estimator. IVW combines SNP-specific Wald ratio estimates in a fixed-effects meta-analysis framework to generate an overall causal effect estimate. UVMR was performed for general frailty and each of the six domain-specific frailty factors (F1-F6), as demonstrated in Figure 1, on two Alzheimer’s disease (AD) GWAS outcome datasets: (1) AD Kunkle ^[17]^ and (2) AD Wightman ^[18]^. In addition, bidirectional MR analyses were performed to test whether genetic liability for AD causally influences frailty.

To assess the robustness of our findings and account for potential pleiotropy, we conducted several sensitivity analyses, including MR-Egger (adjusting for directional pleiotropy), weighted median and penalised weighted median approaches (providing estimates robust to invalid instruments), weighted mode analysis, MR-PRESSO (to detect and correct for outlier SNPs), and leave-one-out analyses to identify influential variants. We also quantified between-SNP heterogeneity using Cochran’s Q and I² statistics to evaluate the consistency of SNP effect estimates across instruments. Multiple testing correction was applied using a false discovery rate (FDR) threshold of *q* < 0.05 based on the number of exposures included in each analysis set. We also calculated the F-statistic for each IV to ensure that we only included strong genetic instruments (F > 10) in our analyses.

#### MR Analyses including Socioeconomic and Longevity/Longevity Controls

We conducted separate multivariable MR (MVMR) for each of the frailty exposures to evaluate whether their associations with AD were independent of educational attainment (EA), income, or longevity. The primary MVMR models included the frailty exposure of interest along with EA and income as simultaneous exposures, allowing us to account for potential socioeconomic confounding. Given evidence that lifespan and mortality risk may act as upstream determinants of both frailty and Alzheimer’s disease, we ran additional MVMR models for each frailty exposure including EA, income, and longevity as simultaneous exposures (Figure 2 displays an example of this MVMR model). This allowed us to test whether frailty – AD associations were robust after adjusting for socioeconomic status and genetic liability to longer life.

**Figure 2.**
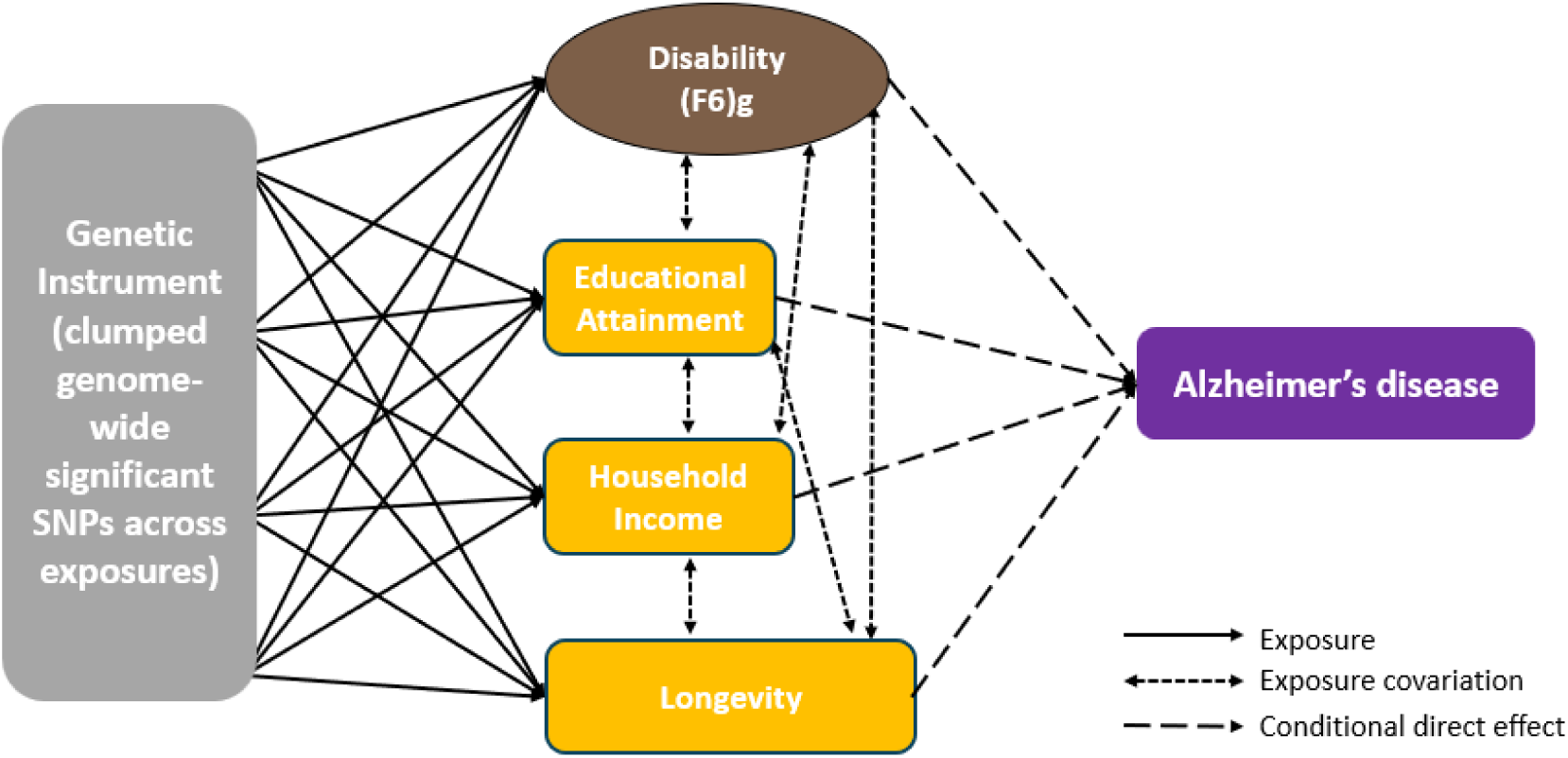
Analytical framework for MVMR analyses of frailty domains and Alzheimer’s disease, accounting for education, income, and longevity. As in Figure 1, we show an example of the model for one frailty factor, but in practice we ran a model like this separately for each of the latent frailty factors.

#### Colocalisation Analyses

To further investigate whether observed MR associations reflected shared causal variants or indirect/pleiotropic pathways, we conducted colocalisation analyses using a Bayesian framework (coloc). These analyses estimate the posterior probability that the same genetic variant influences both the exposure and outcome (PP.H4). Colocalisation was performed as a follow-up for frailty domains showing evidence of association with Alzheimer’s disease (AD) in primary or sensitivity MR analyses, specifically unhealthy lifestyle (F2), poorer cognition (F5), and disability (F6). Regional summary statistics were extracted using ±1 Mb windows around lead SNPs, and analyses were restricted to autosomal regions after exclusion of the APOE locus (chr19:44,909,039–45,912,650; hg19) and the major histocompatibility complex (MHC; chr6:25–34 Mb), due to their complex linkage disequilibrium structure. Posterior probabilities for five hypotheses (H0–H4) were calculated, with particular focus on PP.H4 as evidence for a shared causal variant.

## Results

### Bidirectional UVMR

When the frailty domains were modelled as single exposures, there was no evidence of a significant causal effect of the general frailty factor (GF) or frailty domains linked to limited social support (F1), multimorbidity (F3), metabolic problems (F4), or poorer cognition (F5) on AD (all q > 0.05 – all results shown in Supplementary table 2). In contrast, the unhealthy lifestyle domain frailty (F2) showed a strong and consistent risk-increasing effect across outcomes [AD (Kunkle): OR = 6.60, 95% CI: 2.36-18.50, p = 0.0003, q = 0.002 and AD (Wightman): OR = 3.12, 95% CI: 1.57-6.19, p = 0.001, q = 0.008]. The disability frailty domain (F6) showed a nominally protective effect in AD (Kunkle): OR = 0.56, 95% CI: 0.33-0.97, p = 0.038, but did not survive FDR correction.

Bidirectional MR analyses were conducted using AD genetic liability as the exposure (we tested both AD Kunkle and AD Wightman separately) and the frailty domains (general frailty [GF] and specific domains (F1-F6) as the outcomes (Supplementary Tables 3 and 4). There was no evidence of a causal effect of AD genetic liability on any frailty domains in any dataset (all q > 0.05), indicating no support for a reverse causal relationship between AD risk and genetically predicted frailty traits. This strengthens the interpretation that the significant associations observed in the primary analyses reflect a directional effect of frailty domains on AD risk, rather than reverse causation.

#### Sensitivity and heterogeneity

Instrument strength was adequate (mean F-statistics 32.99-44.86; all >10), with per-SNP F statistics typically >30 (Supplementary Tables 5 and 6). In the multivariable models, conditional F-statistics were markedly lower (Supplementary Table 7), reflecting the shared genetic architecture between frailty, education, income, and longevity. This reduction is expected, as conditional F values in MVMR capture the unique variance in each exposure after accounting for the others, and are therefore not directly comparable to the stronger univariable F-statistics.

Sensitivity analyses using weighted median, weighted mode, and MR-PRESSO were broadly consistent with the IVW results (Supplementary Table 8). The unhealthy lifestyle frailty domain (F2) showed a robust risk-increasing causal effect on AD across methods and datasets. The disability frailty domain (F6) showed a protective causal effect in AD (Kunkle): OR = 0.43, 95% CI: 0.21-0.87, p = 0.019, q = 0.048, that was consistent across sensitivity estimators, although weaker in magnitude. This trend was also observed in the Wightman data, although it was not statistically significant. The poorer cognition frailty domain (F5) also showed evidence of a risk-increasing causal effect in AD (Kunkle) when using the weighted median estimator, indicating that this exposure may have some pleiotropic SNPs included in it. Leave-one-out analyses (Supplementary Table 9) indicated that the unhealthy lifestyle (F2) pathway remained robust across both outcomes, with no single SNP explaining the effect. For the frailty domain disability (F6), the protective causal effect in AD (Kunkle) persisted in leave-one-out analyses, indicating that no single SNP accounted for the association. This trend was also observed for AD (Wightman).

Cochran’s Q tests indicated low-to-moderate heterogeneity for most exposure-outcome pairs (I² < 30%), with notable exceptions for GF-AD (Wightman), F3-AD (Wightman), and especially F5 (poorer cognition), which showed strong heterogeneity across all AD outcomes (I² = 5467%) (Supplementary Tables 10 and 11). Leave-one-out plots did not indicate that removal of any single SNP entirely explained the F5-AD relationships, consistent with heterogeneity being spread across multiple instruments, which was supported by our weighted median and weighted mode estimates for AD (Lambert). However, MR-PRESSO for frailty linked to poorer cognition (F5) identified recurrent outlier variants, including rs67472071 and rs6869910, that contributed disproportionately. rs67472071 lies within the *SPI1* locus, highlighted in the original frailty GWAS as a key locus for F5 and also a well-established AD risk locus ^[33]^.

rs6869910 maps near *PFDN1* and *CYSTM1* and has been implicated in GWAS of intelligence and mathematical ability ^[34]^. The recurrence of these SNPs across outcomes underscores the possibility that the frailty domain poorer cognition captures pleiotropic cognitive variants, reflecting shared genetic architecture with AD rather than an independent pathway.

#### Multivariable MR with education, income, and longevity

The disability frailty domain (F6) showed the clearest evidence of an independent risk increasing effect on AD. In EA + Income-adjusted models, F6 was significant in AD (Wightman): OR = 1.66, 95% CI: 1.19-2.32, p = 0.003 q = 0.009] and when longevity was additionally included, the frailty disability domain with AD (Wightman) effect remained comparable [OR = 1.68, 95% CI: 1.21-2.36, p = 0.002]. These trends were also replicated in the AD (Kunkle) data [OR = 1.52, 95% CI: 0.99-2.35, p = 0.057, q = 0.086, for EA + Income and OR = 1.52, 95% CI: 0.99-2.33, p = 0.057, q = 0.087 for EA + Income + Longevity adjusted models], but were not statistically significant. This pattern of findings indicates that the frailty disability domain is strongly confounded by socioeconomic pathways, but when these are appropriately accounted for, the frailty-specific genetic signal for disability is associated with increased AD risk and this effect is independent of lifespan-related pathways.

The poorer cognition frailty domain (F5) also showed evidence of a risk-increasing effect in AD (Kunkle): β = 0.44, p = 0.008, q = 0.025] in the EA + Income adjusted model, but this attenuated and was no longer significant when longevity was added. This trend was also seen in AD (Wightman), although it was not significant (Supplementary Table 12). This indicates that the UVMR effect estimate was being strongly confounded by socioeconomic-related pathways, but that when these are appropriately adjusted for, there is a strong direct causal effect of frailty-related poorer cognition on AD. However, this effect was attenuated by ∼37% when longevity was included in the model (OR reduced from 1.55 [95% CI: 1.12-2.15] to 1.32 [95% CI: 0.97-1.81]), resulting in a shift from significant to non-significant. This attenuation suggests that lifespan-related pathways may partly account for the observed association, although not fully.

Of particular note, the strong risk-increasing effect of the unhealthy lifestyle frailty domain (F2) seen in the UVMR analysis was completely attenuated to null in the EA + Income adjusted model across both AD datasets, with log-odds reduced by ∼66-94% and ORs falling from ∼3-7 in UVMR to ∼1.1-1.5 in MVMR, with confidence intervals overlapping the null and q-values > 0.05, indicating that this effect is being completely driven via confounding genetic effects of socioeconomic status associated with these behaviours.

All other frailty domains – limited social support (F1), multimorbidity (F3), metabolic problems (F4) and general frailty (GF) – showed no evidence of a causal effect on AD risk after adjustment for education, income, and longevity (all q > 0.05, all ORs close to 1 – full results for all MVMR models are in Supplementary Table 12). Alongside UVMR, and sensitivity results, MVMR results are displayed in Figure 3.

**Figure 3.**
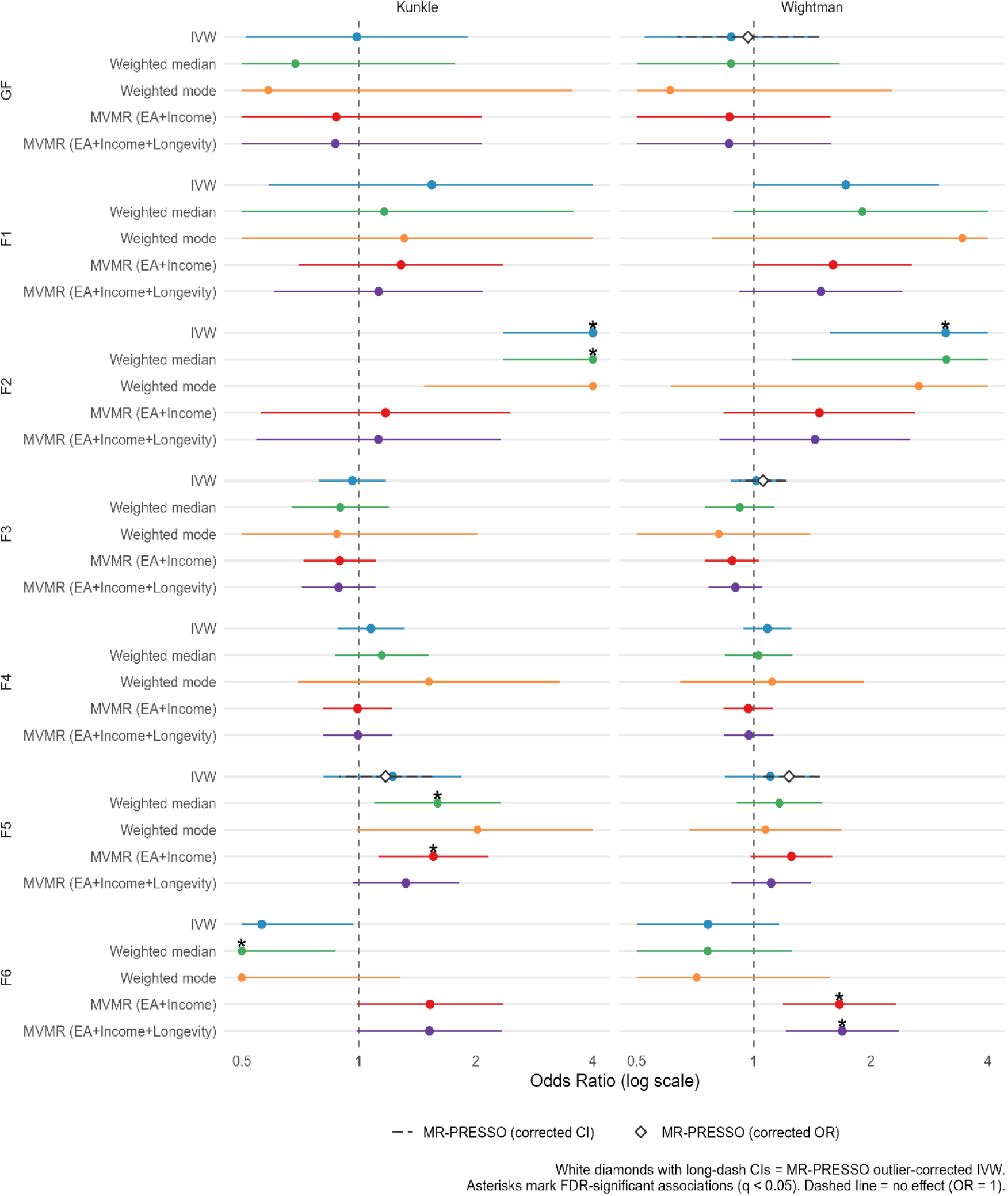
UVMR and MVMR estimates showing the causal effects of genetically derived frailty domains (general frailty [GF] and six domain-specific pathways: limited social support [F1], unhealthy lifestyle [F2], multimorbidity [F3], metabolic problems [F4], poorer cognition [F5], and disability [F6]) on Alzheimer’s disease risk across two GWAS outcome datasets (Kunkle et al., 2019 and Wightman et al., 2021). Points represent beta estimates (log-odds of AD per 1SD increase in frailty), with horizontal lines showing 95% confidence intervals. The dashed vertical line indicates the null (no effect). White diamonds denote MRPRESSO outlier-corrected odds ratios, and asterisks (*) mark associations surviving false discovery rate (FDR) correction.

#### APOE sensitivity analyses

Repeating analyses without removing variants in and around the *APOE* locus (±500 kb) produced consistent findings with the main analyses both in terms of in direction and magnitude of effects. Including *APOE* variants did not change which results survived FDR correction, indicating that the genetic pathways driving the associations between these frailty domains and AD risk are not being disproportionately driven by the *APOE* region. The results of these analyses are included alongside all results in Supplementary Tables 2-12.

#### Downstream colocalisation

Colocalisation analyses provided further insight into the nature of these associations. For the poorer cognition frailty domain (F5), there was moderate evidence of colocalisation with AD at the SPI1 locus (rs67472071), with posterior probabilities for a shared causal variant (PP.H4) of 63.6% in the Kunkle dataset and 59.5% in the Wightman dataset. This locus has been robustly implicated in AD through microglial regulatory pathways, and its recurrence across MR sensitivity analyses and outlier detection supports the presence of shared genetic architecture. In contrast, there was little evidence of colocalisation for the disability frailty domain (F6) or the unhealthy lifestyle domain (F2) after exclusion of the APOE and MHC regions (all PP.H4 < 0.05), suggesting that their observed MR associations are unlikely to be driven by shared causal variants at specific loci. Together, these findings indicate that the F5 signal is likely influenced by pleiotropic AD-related genetic variants, whereas associations observed for F2 and F6 are more consistent with indirect or downstream pathways rather than direct shared genetic effects, colocalisation results are available in Supplementary Table 13.

## Discussion

This study represents the most comprehensive Mendelian randomisation (MR) investigation to date into the potential causal role of frailty in Alzheimer’s disease (AD). By leveraging genetically derived instruments for both general and domain-specific frailty traits, we were able to move beyond traditional single aggregate frailty measures to examine the unique causal relationships between AD and different domains of frailty. Our findings highlight the complexity of frailty as a multidimensional construct, the importance of disaggregating frailty components, and the critical role that socioeconomic and longevity pathways play in frailty and AD risk.

Notably, our univariable MR (UVMR) analyses using genetic instruments derived from a general frailty factor, akin to a frailty index, found no evidence of a causal association with AD. This result aligns with prior null MR findings for the frailty index ^[16]^, suggesting that when frailty is conceptualised as a broad, aggregated measure, its underlying genetic architecture may not capture distinct pathways that causally influence neurodegeneration. Indeed, the general frailty factor from the most recent GWAS frailty study was derived using Genomic SEM to capture shared variance across diverse traits commonly assessed in frailty indices ranging from fatigue and sensory decline to mental health and falls ^[15]^. The null results for this general factor suggest that the accumulation of risk from multiple diverse pathways that contribute to overall frailty is not a direct cause of elevated AD risk. Instead, the observed phenotypic association between frailty and AD is more likely to be due to specific biological mechanisms that affect subcomponents of frailty.

When we used genetic instruments for domain-specific latent frailty factors – each orthogonal to the general factor and capturing genetically distinct subdimensions of frailty – a more nuanced picture emerged. The unhealthy lifestyle frailty domain (F2), which reflects shared genetics between cigarette smoking, arterial stiffness, and physical inactivity, showed strong and consistent associations with increased AD risk across all both AD outcome datasets in UVMR. These associations align with the substantial body of epidemiological evidence linking mid- and late-life smoking, sedentary behaviour, and cardiovascular dysfunction with elevated dementia risk ^[2]^.

However, in our multivariable MR (MVMR) models adjusting for educational attainment and income these effects were substantially attenuated, with effect estimates reduced and no longer statistically significant, echoing recent MR studies suggesting that genetic liability to unhealthy lifestyle factors may operate partly through socioeconomic pathways ^[35]^. This finding reinforces the importance of modelling known confounders when assessing putative risk pathways in MR, and illustrates that while lifestyle behaviours are often conceptualised as modifiable exposures, the genetic predisposition to such behaviours may be partially explained by genetic liability to broader structural determinants such as socioeconomic status. Similarly, there was little evidence of colocalisation between the unhealthy lifestyle frailty domain and AD, further supporting the interpretation that these associations are mediated through broader socioeconomic or behavioural pathways rather than shared genetic loci.

The most striking and consistent effect emerged for the disability frailty domain(F6), a domain encompassing the shared genetics between self-reported disability, poor overall health rating, and eye disorders. In UVMR, the frailty disability domain was associated with a reduced risk of AD, particularly under the weighted median estimator, which survived FDR correction, indicating a protective effect. But this effect became risk-increasing for AD in the MVMR models, when socioeconomic status was adjusted for. This stark reversal in both AD datasets suggests a strong confounding effect of socioeconomic status on the causal relationship between frailty disability pathways and AD. Disability is known to be more prevalent in socioeconomically disadvantaged groups ^[39]^, and gene-environment correlations may amplify this effect. Individuals with genetic predispositions for poor physical health may also encounter environments that worsen these risks, including reduced healthcare access, higher stress, or adverse living conditions. Adjusting for socioeconomic status in multivariable MR may help mitigate this bias by accounting for confounding pathways, allowing the latent biological association between disability-related frailty and AD risk to emerge more clearly.

Consistent with this, colocalisation analyses provided little evidence for shared causal variants between the disability frailty domain and AD after exclusion of APOE and MHC regions, suggesting that the observed associations are unlikely to be driven by locus-specific shared genetic effects.

However, it is also plausible that confounding effects observed in certain frailty domains reflect a form of participation bias in the UK Biobank, where individuals with genotypes linked to both lower socioeconomic status and higher frailty are underrepresented. UK Biobank participants tend to be healthier and more affluent than the general population ^[36]^. Thus, genetic correlations between socioeconomic status and frailty may partly reflect this selection, rather than true shared biological pathways. Previous research, on other traits, has highlighted how the magnitude of genetic effects on intelligence and education varies across socioeconomic contexts, with indirect genetic effects from parental genotypes shaping the rearing environment ^[37]^. These context-dependent effects mean that genetic correlations between socioeconomic-linked traits and health outcomes can be inflated or distorted in selected samples such as UK Biobank. Similarly, other research found that only around one quarter of the polygenic signal for income reflects direct genetic effects, with the remainder likely reflecting environmental pathways via education and broader socioeconomic exposures ^[38]^. Together, these findings suggest that socioeconomic-linked genetic variants capture a complex blend of direct and indirect effects, which may obscure the true biological relationship between disability and AD risk in unadjusted univariable MR models.

Beyond socioeconomic confounding, other forms of selection bias may also influence these results. Survival bias is one possibility, including right censoring – where individuals with higher genetic liability to disability die earlier, reducing the probability of surviving to ages at which AD is typically diagnosed, and thereby creating an apparent protective effect in unadjusted analyses ^[40,41]^. To examine this possibility, we ran sensitivity MVMR models including longevity alongside educational attainment and household income. These analyses slightly attenuated the F6-AD relationship, but did not materially alter the main findings, suggesting that survival bias is unlikely to impact the observed effect. Nevertheless, the role of longevity is conceptually complex: while accounting for survival can reduce bias from early mortality, it may also remove part of the causal pathway through which disability-related frailty influences AD risk since frailty and longevity are inextricably linked, and likely to be highly negatively collinear. Our findings should therefore be interpreted with this caveat in mind.

Previous MR studies have shown that socioeconomic traits – including education, income, and social isolation – causally influence frailty ^[42]^. Moreover, transcriptomic evidence demonstrates that SES-based inequalities in molecular pathways relevant to ageing and chronic disease are already detectable by early adulthood ^[43]^. These findings underscore the complexity of disentangling social, biological, and genetic pathways in the aetiology of AD.

In contrast, the poorer cognition frailty domain (F5) showed a different conditional pattern. Weighted median UVMR identified a risk-increasing effect for F5 on AD (Kunkle), surviving FDR correction, and this signal re-emerged in MVMR when adjusting for education and income. However, unlike the frailty disability domain, the frailty domain linked to poorer cognition was reduced once longevity was included in the MVMR, suggesting that this association with AD is strongly mediated or shared with lifespan-related genetic pathways. Furthermore, heterogeneity between the genetic variants for the frailty domain, which represents poorer cognition, was high, and MR-PRESSO flagged rs67472071, a SNP located within the *SPI1* locus, as a recurrent outlier across outcomes. *SPI1* encodes the transcription factor PU.1, a key regulator of microglial function, that has been robustly implicated in AD pathogenesis ^[33]^. This interpretation is consistent with the Genomic SEM frailty GWAS, which identified this frailty domain as the only one exhibiting significant genetic overlap with AD, implicating loci such as SPI1 – a well-established AD risk region ^[15,33]^. The recurrence of this locus suggests that the poorer cognition signal may partly reflect directly shared pleiotropic effects of established AD risk variants, rather than an independent causal pathway from frailty to AD. This interpretation is supported by colocalisation analyses, which demonstrated moderate evidence for a shared causal variant at the SPI1 locus (rs67472071; PP.H4 ≈ 60– 64% across AD datasets), reinforcing that the observed association is likely driven by shared genetic architecture rather than a distinct causal effect of frailty on AD.

It is important to note that our MR analyses estimate the causal effects of lifelong genetic liability to frailty domains, rather than measured levels at a specific age. Although the frailty indicators used to derive genetic instruments were assessed in mid-to late-life in UK Biobank, the associated variants capture vulnerability to these traits across the life course. For example, one disability item was based on genetic variants associated with the UK Biobank question “Do you have any long-standing illness, disability or infirmity?”. While phenotypically broad, this signal likely indexes liability to physical impairment that may emerge earlier or later in life. The persistence of its causal effect on AD risk, even after adjustment for SES, suggests a potential pathway linking early or midlife health deficits to later neurodegeneration. From a prevention perspective, this highlights that frailty and disability measures collected in midlife could serve as risk markers for dementia, supporting earlier identification and intervention before neurodegenerative processes become established.

The variability in results across the Kunkle ^[17]^ and Wightman ^[18]^ outcome datasets highlight how sample composition, diagnostic methodology, and cohort structure can affect MR estimates. Kunkle et al., 2019 provides a gold-standard case-control design, with clinically or autopsy-confirmed diagnoses and close age-matching between cases and controls, minimising confounding by age-related comorbidity. Wightman et al., 2021, in contrast, offers greater statistical power as the largest clinically defined case-control GWAS to date, but its reliance on population-based biobank cohorts and limited reporting on age-matching complicate interpretation. By analysing both datasets, we balanced diagnostic precision (Kunkle) with statistical power (Wightman), enabling triangulation of causal estimates across complementary designs. This approach helps to explain why some effects were more evident in Kunkle, while socioeconomic mediation effects appeared stronger in Wightman, reflecting differences in case ascertainment and cohort structure.

Finally, our bidirectional MR analyses revealed no evidence of reverse causation from AD liability to any of the frailty domains outcome datasets. This consistent absence of reverse associations strengthens the argument for a unidirectional causal pathway from frailty to AD. Our genetically informed approach supports the phenotypic interpretation ^[8]^: genetically proxied frailty-particularly in domains like disability appears to predispose individuals to AD rather than result from it.

Our findings emphasise the value of genetic instruments in disaggregating the multidimensional construct of frailty. While the general frailty factor mirrors traditional frailty instruments in showing no causal effect, the genetically isolated subdimensions relating to frailty – particularly those linked to lifestyle, metabolic function, and disability – reveal distinct, biologically and socially mediated pathways to AD. These results suggest that frailty is not a unitary risk factor for Alzheimer’s disease but a complex constellation of genetically separable traits with differential implications for prevention. Future research should prioritise replication of these findings, expanded to other dementia subtypes, in ancestrally diverse samples, which may experience different interactions between genetic and environmental contributions to frailty. It will also be important to explore the mechanisms by which genetically encoded disability vulnerability contributes to neurodegeneration.

## Supporting information

Supplementary_Materials_Domains of Frailty as Early Risk Factors for Alzheimer's disease

Supplementary_Tables_Domains of Frailty as Early Risk Factors for Alzheimer's disease

## Data Availability

Availability of data and material: Data is all publicly available via https://www.ebi.ac.uk/gwas/
. Study accession codes for AD studies: Kunkle et al. – GCST007511, Wightman et al. – GCST013196. Study accession codes for Frailty Domains – GCST90624046–GCST90624053. Study accession codes for confounders: Income – GCST009523, and Longevity – GCST006697. With the exception of the EA summary statistics, which are available by creating an account with the Social Science Genetic Association Consortium (https://www.thessgac.org/
) and downloading the summary statistics for PMID: 30038396. Details for all summary statistics used are available in Supplementary Table 1.

## Acknowledgments

Concept and Design: JP Flint, IF Foote, SR Cox, M Luciano, MK Lupton, Data analysis: JP Flint; Drafting of the manuscript: JP Flint; Critical revision of the manuscript: JP Flint, IF Foote, MK Lupton, SR Cox, M Luciano, TC Russ, A Marshall, R Rahman, David D Ward; This research was funded by the Legal & General Group (research grant to establish the independent Advanced Care Research Centre at the University of Edinburgh). The funder had no role in the conduct of the study, interpretation, or the decision to submit for publication. The views expressed are those of the authors and not necessarily those of legal and general. The authors declare no competing interests.

## Availability of data and material

Data is all publicly available via https://www.ebi.ac.uk/gwas/. Study accession codes for AD studies: Kunkle et al. – GCST007511, Wightman et al. – GCST013196. Study accession codes for Frailty Domains – GCST90624046-GCST90624053. Study accession codes for confounders: Income – GCST009523, and Longevity – GCST006697. With the exception of the EA summary statistics, which are available by creating an account with the Social Science Genetic Association Consortium (https://www.thessgac.org/) and downloading the summary statistics for PMID: 30038396. Details for all summary statistics used are available in Supplementary Table 1.

## References

1. Deuschl G, Beghi E, Fazekas F, Varga T, Christoforidi KA, Sipido E, Bassetti CL, Vos T, Feigin VL. The burden of neurological diseases in Europe: an analysis for the Global Burden of Disease Study 2017. The Lancet Public Health. 2020 Oct 1;5(10):e551–67

2. Nichols E, Vos T. The estimation of the global prevalence of dementia from 1990-2019 and forecasted prevalence through 2050: an analysis for the Global Burden of Disease (GBD) study 2019. Alzheimer’s & Dementia. 2021 Dec;17:e051496.

3. Livingston G, Huntley J, Liu KY, Costafreda SG, Selbæk G, Alladi S, Ames D, Banerjee S, Burns A, Brayne C, Fox NC. Dementia prevention, intervention, and care: 2024 report of the Lancet standing Commission. The Lancet. 2024 Aug 10;404(10452):572–628.

4. Sanderson E, Glymour MM, Holmes MV, Kang H, Morrison J, Munafò MR, Palmer T, Schooling CM, Wallace C, Zhao Q, Davey Smith G. Mendelian randomization. Nature reviews Methods primers. 2022 Feb 10;2(1):6.

5. Thorp JG, Mitchell BL, Gerring ZF, Ong JS, Gharahkhani P, Derks EM, et al. Genetic evidence that the causal association of educational attainment with reduced risk of - Alzheimer dementiais driven by intelligence. Neurobiol Aging. 2022 Nov;119:127–35.

6. Clegg A, Young J, Iliffe S, Rikkert MO, Rockwood K. Frailty in elderly people. Lancet. 2013 Mar 2;381(9868):752–62.

7. Kim DH, Rockwood K. Frailty in Older Adults. N Engl J Med. 2024 Aug 8;391(6):538–48.

8. Ward DD, Flint JP, Littlejohns TJ, Foote IF, Canevelli M, Wallace LMK, et al. Frailty Trajectories Preceding Dementia in the US and UK. JAMA Neurol. 2025 Jan 1;82(1):61–71.

9. Petermann-Rocha F, Lyall DM, Gray SR, Esteban-Cornejo I, Quinn TJ, Ho FK, Pell JP, CelisMorales C. Associations between physical frailty and dementia incidence: a prospective study from UK Biobank. The Lancet Healthy Longevity. 2020 Nov 1;1(2):e58–68.

10. Ruggiero C. Understanding Aging, Frailty, and Resilience. InThe Frail Surgical Patient: A Geriatric Approach Beyond Age 2025 Jan 28 (pp. 49–66). Cham: Springer Nature Switzerland.

11. Atkins JL, Jylhävä J, Pedersen NL, Magnusson PK, Lu Y, Wang Y, Hägg S, Melzer D, Williams DM, Pilling LC. A genome-wide association study of the frailty index highlights brain pathways in ageing. Aging cell. 2021 Sep;20(9):e13459.

12. Ye Y, Noche RB, Szejko N, Both CP, Acosta JN, Leasure AC, Brown SC, Sheth KN, Gill TM, Zhao H, Falcone GJ. A genome-wide association study of frailty identifies significant genetic correlation with neuropsychiatric, cardiovascular, and inflammation pathways. Geroscience. 2023 Aug;45(4):2511–23.

13. Mak JK, Qin C, Krüger M, Kuukka A, Hägg S, Lin J, Jylhävä J. Large-scale genome-wide analyses with proteomics integration reveal novel loci and biological insights into frailty. Nature Aging. 2025 Aug 5:1–2.

14. Pridham G, Rockwood K, Rutenberg A. Efficient representations of binarized health deficit data: the frailty index and beyond. Geroscience. 2023 Jun;45(3):1687–711.

15. Foote IF, Flint JP, Fürtjes AE, Lawrence JM, Mullin DS, Fisk JD, Karakach TK, Rutenberg A, Martin NG, Lupton MK, Llewellyn DJ. Uncovering the multivariate genetic architecture of frailty with genomic structural equation modeling. Nature Genetics. 2025 Aug 4:1–2.

16. Enduru N, Fernandes BS, Zhao Z. Dissecting the shared genetic architecture between Alzheimer dementia and frailty: a cross-trait meta-analyses of genome-wide association studies. Front Genet. 2024 Apr 19;15:1376050.

17. Kunkle BW, Grenier-Boley B, Sims R, Bis JC, Damotte V, Naj AC, et al. Genetic meta-analysis of diagnosed -Alzheimer dementia identifies new risk loci and implicates Aβ, tau, immunity and lipid processing. Nat Genet. 2019 Mar;51(3):414–30.

18. Wightman DP, Jansen IE, Savage JE, Shadrin AA, Bahrami S, Holland D, et al. A genome-wide association study with 1,126,563 individuals identifies new risk loci for Alzheimer’s disease. Nat Genet. 2021 Sep;53(9):1276–82.

19. Grant AJ, Burgess S. Pleiotropy robust methods for multivariable Mendelian randomization. Stat Med. 2021 Nov 20;40(26):5813–30.

20. Wu Y, Sun Z, Zheng Q, Miao J, Dorn S, Mukherjee S, Fletcher JM, Lu Q. Pervasive biases in proxy genome-wide association studies based on parental history of Alzheimer’s disease. Nature genetics. 2024 Dec;56(12):2696–703.

21. Escott-Price V, Hardy J. Genome-wide association studies for Alzheimer’s disease: bigger is not always better. Brain Communications. 2022 Jun 1;4(3):fcac125.

22. Lee JJ, Wedow R, Okbay A, Kong E, Maghzian O, Zacher M, et al. Gene discovery and polygenic prediction from a genome-wide association study of educational attainment in 1.1 million individuals. Nat Genet. 2018 Jul 23;50(8):1112–21.

23. Hill WD, Davies NM, Ritchie SJ, Skene NG, Bryois J, Bell S, et al. Genome-wide analysis identifies molecular systems and 149 genetic loci associated with income. Nat Commun. 2019 Dec 16;10(1):5741.

24. Pilling LC, Kuo CL, Sicinski K, Tamosauskaite J, Kuchel GA, Harries LW, Herd P, Wallace R, Ferrucci L, Melzer D. Human longevity: 25 genetic loci associated in 389,166 UK biobank participants. Aging (Albany NY). 2017 Dec 6;9(12):2504.

25. Mez J, Marden JR, Mukherjee S, Walter S, Gibbons LE, Gross AL, Zahodne LB, Gilsanz P, Brewster P, Nho K, Crane PK. Alzheimer’s disease genetic risk variants beyond APOE ε4 predict mortality. Alzheimer’s & Dementia: Diagnosis, Assessment & Disease Monitoring. 2017 Jan 1;8:188–95.

26. Sanderson E, Glymour MM, Holmes MV, Kang H, Morrison J, Munafò MR, et al. Mendelian randomization. Nat Rev Methods Primers [Internet]. 2022 Feb 10;2(1). Available from: 10.1038/s43586-021-00092-5

27. Zheng J, Baird D, Borges MC, Bowden J, Hemani G, Haycock P, et al. Recent developments in Mendelian randomization studies. Curr Epidemiol Rep. 2017 Nov 22;4(4):330–45.

28. Skrivankova VW, Richmond RC, Woolf BA, Yarmolinsky J, Davies NM, Swanson SA, VanderWeele TJ, Higgins JP, Timpson NJ, Dimou N, Langenberg C. Strengthening the reporting of observational studies in epidemiology using Mendelian randomization: the STROBE-MR statement. Jama. 2021 Oct 26;326(16):1614–21.

29. A Language and Environment for Statistical Computing_. R Foundation for Statistical Computing. A Language and Environment for Statistical Computing_ R Foundation for Statistical Computing.

30. Hemani G, Zheng J, Elsworth B, Wade KH, Haberland V, Baird D, et al. The MR-Base platform supports systematic causal inference across the human phenome. Elife [Internet]. 2018 May 30;7. Available from: 10.7554/eLife.34408

31. Sanderson E, Spiller W, Bowden J. Testing and correcting for weak and pleiotropic instruments in two-sample multivariable Mendelian randomization. Stat Med. 2021 Nov 10;40(25):5434–52.

32. Verbanck M, Chen CY, Neale B, Do R. Detection of widespread horizontal pleiotropy in causal relationships inferred from Mendelian randomization between complex traits and diseases. Nat Genet. 2018 May;50(5):693–8.

33. Bellou E, Escott-Price V. Are Alzheimer’s and coronary artery diseases genetically related to longevity?. Frontiers in Psychiatry. 2023 Jan 6;13:1102347.

34. Zhang Z, Zhu Y, Zhang J, He W, Han C. Identification of novel proteins associated with intelligence by integrating genome-wide association data and human brain proteomics. Plos one. 2025 Feb 21;20(2):e0319278.

35. Meng X, Li X, Cao M, Dong J, Wang H, Cao W, Liu D, Wang Y. Summarizing attributable factors and evaluating risk of bias of Mendelian randomization studies for Alzheimer’s dementia and cognitive status: a systematic review and meta-analysis. Systematic Reviews. 2025 Mar 13;14(1):61.

36. Fry A, Littlejohns TJ, Sudlow C, Doherty N, Adamska L, Sprosen T, Collins R, Allen NE. Comparison of sociodemographic and health-related characteristics of UK Biobank participants with those of the general population. American journal of epidemiology. 2017 Nov 1;186(9):1026–34.

37. Hill WD. Environmental Influences on Genetic Contributions to Intelligence and Education. American Journal of Psychiatry. 2021 Jul;178(7):582–3.

38. Kweon H, Burik CA, Ning Y, Ahlskog R, Xia C, Abner E, Bao Y, Bhatta L, Faquih TO, de Feijter M, Fisher P. Associations between common genetic variants and income provide insights about the socio-economic health gradient. Nature human behaviour. 2025 Apr;9(4):794–805.

39. Yip JL, Muthy Z, Peto T, Lotery A, Foster PJ, Patel P. Socioeconomic risk factors and agerelated macular degeneration in the UK Biobank study. BMJ open ophthalmology. 2021 Feb 1;6(1):e000585.

40. Coemans M, Verbeke G, Döhler B, Süsal C, Naesens M. Bias by censoring for competing events in survival analysis. bmj. 2022 Sep 13;378.

41. Rojas-Saunero LP, Young JG, Didelez V, Ikram MA, Swanson SA. Considering questions before methods in dementia research with competing events and causal goals. American Journal of Epidemiology. 2023 Aug;192(8):1415–23.

42. Huang J, Gui Y, Wu J, Xie Y. Causal effects of socioeconomic traits on frailty: a Mendelian randomization study. Front Med (Lausanne). 2024 Jul 12;11:1344217.

43. Shanahan MJ, Cole SW, Ravi S, Chumbley J, Xu W, Potente C, et al. Socioeconomic inequalities in molecular risk for chronic diseases observed in young adulthood. Proc Natl Acad Sci U S A. 2022 Oct 25;119(43):e2103088119.

