## Supplementary_Materials_Domains of Frailty as Early Risk Factors for Alzheimer's disease for "Domains of Frailty as Early Risk Factors for Alzheimer’s Disease – Genetic and Causal Evidence"

**STROBE-MR checklist of recommended items to address in reports of Mendelian randomization studies**[1 2]

| Item No. | Section | Checklist item | Page No. | Relevant text from manuscript |
|---|---|---|---|---|
| 1 | **TITLE and ABSTRACT** | Indicate Mendelian randomization (MR) as the study's design in the title and/or the abstract if that is a main purpose of the study | p. 2 | Title includes 'Genetic and Causal Evidence' whilst abstract states: 'We applied a two-stage Mendelian randomisation (MR) framework...' |
| | **INTRODUCTION** | | | |
| 2 | **Background** | Explain the scientific background and rationale for the reported study. What is the exposure? Is a potential causal relationship between exposure and outcome plausible? Justify why MR is a helpful method to address the study question | p. 3 | "Frailty has consistently been associated with increased risk of cognitive decline and AD [6,7]. In a recent phenotypic analysis of ~30,000 participants across four UK and US cohorts, it was found that frailty levels were elevated 8 to 20 years prior to AD onset, with an acceleration in health decline 4 to 9 years before diagnosis [8]." |
| 3 | **Objectives** | State specific objectives clearly, including pre-specified causal hypotheses (if any). State that MR is a method that, under specific assumptions, intends to h causal effects | p. 4 and p.3 and 8 | "we used a bidirectional two-stage MR approach to investigate whether genetically derived frailty sub-domains have a causal impact on AD. First, we performed univariable MR (UVMR) to estimate the independent effects of general frailty and each of the six domain- |

specific factors on AD risk, using summary statistics from two large GWAS of clinically diagnosed AD. We focused on Kunkle et al. (2019) and Wightman et al. (2021), which together provide complementary strengths: rigorous diagnostic ascertainment with close age-matching (Kunkle et al. 2019) and the largest case–control sample to date through integration of biobank and clinical cohorts (Wightman et al. 2021, excluding proxy cases) [17-18]. We then used multivariable MR (MVMR) to assess whether frailty exerts a direct causal effect on AD, or whether its influence is mediated or confounded by socioeconomic factors, lifespan-related selection, or other indirect pathways [19]. Our study provides the first comprehensive MR investigation of frailty and AD by leveraging a multivariate genetic framework to offer a more nuanced understanding of how specific aspects of frailty may contribute to AD risk, whilst adhering to the STROBE-MR reporting guidelines to ensure methodological transparency and reproducibility."

MR and assumptions are also explained in detail on pages 3 and 8.

| | | | | |
|---|---|---|---|---|
| | **METHODS** | | | |
| 4 | **Study design and data sources** | Present key elements of the study design early in the article. Consider including a table listing sources of data for all phases of the study. For each data source contributing to the analysis, describe the following: | p.7 and supplementary | Table 1 includes a condensed table of all GWAS summary statistics in the MR analyses. Supplementary Table 1 then gives further detail. |
| | | a) Setting: Describe the study design and the underlying population, if possible. Describe the setting, locations, and relevant dates, including periods of recruitment, exposure, follow-up, and data collection, when available. | | |
| | | b) Participants: Give the eligibility criteria, and the sources and methods of selection of participants. Report the sample size, and whether any power or sample size calculations were carried out prior to the main analysis | | |
| | | c) Describe measurement, quality control and selection of genetic variants | | |
| | | d) For each exposure, outcome, and other relevant variables, describe methods of assessment and diagnostic criteria for diseases | | |
| | | e) Provide details of ethics committee approval and participant informed consent, if relevant | | |
| 5 | **Assumptions** | Explicitly state the three core IV assumptions for the main analysis (relevance, independence and exclusion restriction) as well assumptions for any additional or sensitivity analysis | p. 8 | Methods: “Mendelian randomisation (MR) is a statistical method that uses genetic variants as instrumental variables (IVs) to test for causal relationships between an exposure and an outcome [26,27]. For a genetic variant to be considered a valid instrument, three core assumptions must be satisfied: (1) the variant is robustly associated with the exposure (relevance), (2) it affects the outcome only via the exposure (exclusion restriction), and (3) it is not associated with confounders of the |

| | | | | |
|---|---|---|---|---|
| | | | | exposure–outcome relationship (independence).” |
| 6 | **Statistical methods: main analysis** | Describe statistical methods and statistics used | p. 8-10 | IVW as main estimator; MR-Egger, weighted median/mode as sensitivity. SNPs clumped ($r^2$<0.001, 10,000 kb). Exposures scaled per 1-SD. Multiple testing: FDR. Missing data: not applicable. Figures also given to display the UVMR and MVMR |
| | a) | Describe how quantitative variables were handled in the analyses (i.e., scale, units, model) | | |
| | b) | Describe how genetic variants were handled in the analyses and, if applicable, how their weights were selected | | |
| | c) | Describe the MR estimator (e.g. two-stage least squares, Wald ratio) and related statistics. Detail the included covariates and, in case of two-sample MR, whether the same covariate set was used for adjustment in the two samples | | |
| | d) | Explain how missing data were addressed | | |
| | e) | If applicable, indicate how multiple testing was addressed | | |
| 7 | **Assessment of assumptions** | Describe any methods or prior knowledge used to assess the assumptions or justify their validity | p. 11-12 and supplementary | Assessment of assumptions using MR-Egger intercept, MR-PRESSO, Cochran’s Q, $I^2$, leave-one-out. All given in supplementary tables and in text. |
| 8 | **Sensitivity analyses and additional analyses** | Describe any sensitivity analyses or additional analyses performed (e.g. comparison of effect estimates from different approaches, independent replication, bias analytic techniques, validation of instruments, simulations) | pp. 11-12 and supplementary | Sensitivity analyses: MR-Egger, weighted median, weighted mode, MR-PRESSO, leave-one-out, bidirectional MR. |
| 9 | **Software and pre-registration** | | p. 8 | Software: 'All MR analyses were conducted in R (version 4.3.1) using |

| | | | | |
|---|---|---|---|---|
| | | | | TwoSampleMR, MVMR, MR-PRESSO.' Pre-registration: not reported. Suggested: 'The analysis was not pre-registered.' |
| | | a) Name statistical software and package(s), including version and settings used | | |
| | | b) State whether the study protocol and details were pre-registered (as well as when and where) | | |
| | **RESULTS** | | | |
| 10 | **Descriptive data** | | p. 7 | Table 1 and supplementary Table 1 |
| | | a) Report the numbers of individuals at each stage of included studies and reasons for exclusion. Consider use of a flow diagram | | |
| | | b) Report summary statistics for phenotypic exposure(s), outcome(s), and other relevant variables (e.g. means, SDs, proportions) | | |
| | | c) If the data sources include meta-analyses of previous studies, provide the assessments of heterogeneity across these studies | | |
| | | d) For two-sample MR:<br>i. Provide justification of the similarity of the genetic variant-exposure associations between the exposure and outcome samples<br>ii. Provide information on the number of individuals who overlap between the exposure and outcome studies | | |
| 11 | **Main results** | | p. 11-13 | |
| | | a) Report the associations between genetic variant and exposure, and between genetic variant and outcome, preferably on an interpretable scale | | |
| | | b) Report MR estimates of the relationship between exposure and outcome, and the measures of uncertainty from the MR analysis, on an interpretable scale, such as odds ratio or relative risk per SD difference | | |
| | | c) If relevant, consider translating estimates of relative risk into absolute risk for a meaningful time period | | |

| | | | | |
|---|---|---|---|---|
| | | d) Consider plots to visualize results (e.g. forest plot, scatterplot of associations between genetic variants and outcome versus between genetic variants and exposure) | | |
| 12 | **Assessment of assumptions** | | p.11 and supplementary tables 1,2 and 3. | Results: 'Leave-one-out analyses… no single variant driving findings. Cochran's Q indicated little heterogeneity for most.' |
| | | a) Report the assessment of the validity of the assumptions | | |
| | | b) Report any additional statistics (e.g., assessments of heterogeneity across genetic variants, such as $I^2$, Q statistic or E-value) | | |
| 13 | **Sensitivity analyses and additional analyses** | | p. 11-12 | Results: Sensitivity analyses (MR-PRESSO, leave-one-out). Bidirectional MR: no evidence of reverse causation. |
| | | a) Report any sensitivity analyses to assess the robustness of the main results to violations of the assumptions | | |
| | | b) Report results from other sensitivity analyses or additional analyses | | |
| | | c) Report any assessment of direction of causal relationship (e.g., bidirectional MR) | | |
| | | d) When relevant, report and compare with estimates from non-MR analyses | | |
| | | e) Consider additional plots to visualize results (e.g., leave-one-out analyses) | | |
| | **DISCUSSION** | | | |
| 14 | **Key results** | Summarize key results with reference to study objectives | p. 14 | Discussion: 'Notably, GF null; F2 strong associations but attenuated with SES confounding. F5 and F6 emerged as casual exposures in MVMR, with F6 (pathways linked to disability) showing a direct effect after control for SES and longevity. |

| | | | | |
|---|---|---|---|---|
| 15 | **Limitations** | Discuss limitations of the study, taking into account the validity of the IV assumptions, other sources of potential bias, and imprecision. Discuss both direction and magnitude of any potential bias and any efforts to address them | p. 15–16 | Discussion: Limitations include: UK Biobank participation bias, survival bias, heterogeneity across GWAS datasets. |
| 16 | **Interpretation** | | p. 14–17 | Discussion: Meaning: frailty is multidimensional. Mechanisms: SES pathways, disability biology, gene–environment correlations. Clinical: disability as early-life/midlife dementia risk marker. |
| | | a) Meaning: Give a cautious overall interpretation of results in the context of their limitations and in comparison with other studies | | |
| | | b) Mechanism: Discuss underlying biological mechanisms that could drive a potential causal relationship between the investigated exposure and the outcome, and whether the gene-environment equivalence assumption is reasonable. Use causal language carefully, clarifying that IV estimates may provide causal effects only under certain assumptions | | |
| | | c) Clinical relevance: Discuss whether the results have clinical or public policy relevance, and to what extent they inform effect sizes of possible interventions | | |
| 17 | **Generalizability** | Discuss the generalizability of the study results (a) to other populations, (b) across other exposure periods/timings, and (c) across other levels of exposure | p. 17 | Discussion: 'Future research should prioritise replication in ancestrally diverse samples and across other dementia subtypes.' |
| | **OTHER INFORMATION** | | | |
| 18 | **Funding** | Describe sources of funding and the role of funders in the present study and, if applicable, sources of funding for the databases and original study or studies on which the present study is based | p.1 | Funding: If absent, suggested: 'This work was supported by [funders]. Funders had no role in study design, analysis, or interpretation.' |

| | | | | |
|---|---|---|---|---|
| 19 | **Data and data sharing** | Provide the data used to perform all analyses or report where and how the data can be accessed, and reference these sources in the article. Provide the statistical code needed to reproduce the results in the article, or report whether the code is publicly accessible and if so, where | p. 1, 5, 6 (Table 1) | GWAS data are public (Foote et al. 2025, Kunkle 2019, Wightman 2021). Suggested: 'All GWAS data are publicly available; analysis scripts are available upon request.'<br><br>“Availability of data and material: Data is all publicly available via https://www.ebi.ac.uk/gwas/ Study accession codes for AD studies: Kunkle et al. - GCST007511, Wightman et al. - GCST013196. Study accession codes for Frailty Factors - GCST90624046–GCST90624053, Study accession codes for confounders: Income - GCST009523, and Longevity - GCST006697. With the exception of the EA summary statistics, which are available by creating an account with the Social Science Genetic Association Consortium (https://www.thessgac.org/) and downloading the summary statistics for PMID: 30038396. Details for all summary statistics used are available in Supplementary Table 1.” |
| 20 | **Conflicts of Interest** | All authors should declare all potential conflicts of interest | p.1 | The authors declare no competing interests. |

This checklist is copyrighted by the Equator Network under the Creative Commons Attribution 3.0 Unported (CC BY 3.0) license.

1. Skrivankova VW, Richmond RC, Woolf BAR, Yarmolinsky J, Davies NM, Swanson SA, et al. Strengthening the Reporting of Observational Studies in Epidemiology using Mendelian Randomization (STROBE-MR) Statement. JAMA. 2021;under review.

2. Skrivankova VW, Richmond RC, Woolf BAR, Davies NM, Swanson SA, VanderWeele TJ, et al. Strengthening the Reporting of Observational Studies in Epidemiology using Mendelian Randomisation (STROBE-MR): Explanation and Elaboration. BMJ. 2021;375:n2233.
